# Excess Serum Interleukin-18 Distinguishes Patients with Pathogenic Mutations in *PSTPIP1*

**DOI:** 10.1101/2021.02.22.21251857

**Authors:** Deborah L. Stone, Amanda Ombrello, Juan I. Arostegui, Corinne Schneider, Adriana de Jesus, Charlotte Girard-Guyonvarc’h, Cem Gabay, Wonyong Lee, Jae Jin Chae, Ivona Aksentijevich, Raphaela Goldbach-Mansky, Daniel L. Kastner, Scott W. Canna

**Affiliations:** Inflammatory Disease Section, Intramural Research Program, NHGRI; Department of Immunology, Hospital Clinic; Institut d’Investigacions Biomédiques August Pi i Sunyer, Barcelona, Spain; University of Pittsburgh/UPMC Children’s Hospital of Pittsburgh; Translational Autoinflammatory Diseases Section (TADS), Laboratory of Clinical Immunology and Microbiology, Intramural Research Program, NIAID; Division of Rheumatology, Department of Medicine, University Hospital of Geneva and Faculty of Medicine University of Geneva, Geneva, Switzerland

**Author notes:** Address Correspondence to: Scott W. Canna, 8124 Rangos Bldg, 530 45^th^ St, Pittsburgh, PA 15201.

## Abstract

**Objective:** Dominantly-inherited mutations in *PSTPIP1* cause a family of monogenic autoinflammatory diseases epitomized by Pyogenic Arthritis, Pyoderma gangrenosum, and Acne (PAPA) syndrome. The connections between PSTPIP1 and PAPA are poorly understood, although *in vitro* evidence suggests increased activation of the pyrin-inflammasome. We sought to identify biomarkers of potential mechanistic, diagnostic, and therapeutic utility specific to autoinflammatory diseases.

**Methods:** Clinical and genetic data and sera were obtained from patients referred with concern for PAPA syndrome, as well as relevant disease controls. Serum Interleukin-18 (IL-18) and related biomarkers were assessed by bead-based assay.

**Results:** Symptoms in *PSTPIP1* mutation-positive PAPA patients overlapped with those of mutation-negative PAPA-like patients, but the former were younger at onset and had more arthritis. We found uniform elevation of total IL-18 in PAPA patients at a level approaching NLRC4-associated Macrophage Activation Syndrome (MAS) and well beyond Familial Mediterranean Fever. IL-18 elevation in PAPA patients’ sera persisted despite fluctuations in disease activity. IL-18 Binding Protein (IL-18BP) was modestly elevated, and as such PAPA patients had detectable free IL-18. PAPA patients did not develop MAS, and CXCL9 (an indicator of Interferon-gamma activity) was rarely elevated in their sera.

**Conclusion:** PAPA syndrome is a refractory, and often disabling monogenic autoinflammatory disease associated with chronic elevation of serum IL-18, but not risk for MAS. This finding instructs our understanding of the origins of excess IL-18, its potential spectrum of pathogenic effects, and the possible role for IL-18 blockade in refractory PAPA syndrome.

## INTRODUCTION

Pyogenic arthritis, pyoderma gangrenosum, and acne (PAPA) syndrome was first associated with dominant mutations in *PSTPIP1* in 2002^1^, but since that time the links between such mutations and the protean clinical manifestations have been challenging to unravel. *PSTPIP1* mutations are now known to represent an array of manifestations that includes systemic features, early onset (sterile) neutrophilic arthritis, and variable cutaneous involvement. Skin manifestations include simple ulceration, pyoderma gangrenosum, cystic acne, and hidradenitis suppurativa^2, 3^. Other clinical findings can include cytopenias, lymphadenopathy, and hepatosplenomegaly, particularly in the context of high serum Zinc and extremely elevated S100 proteins^4^. This spectrum has been collectively termed *PSTPIP1*-associated inflammatory diseases (PAID)^3^. Causative mutations operate in a dominant inheritance pattern, and include both missense mutations and potentially promotor microsatellite expansions^2, 5^.

Mechanistically, several *PSTPIP1* mutations have been shown to increase binding to pyrin, the protein mutated in Familial Mediterranean Fever (FMF) and activate both the pyrin-and NLRP3-inflammasomes. PSTPIP1 also interacts with the actin cytoskeleton and regulates the Wiskott-Aldrich Syndrome Protein, and *PSTPIP1* mutants potentially alter actin dynamics and thereby pyrin-inflammasome activation and innate immune cell motility^6, 7^.

Interleukin-18 (IL-18) is expressed by macrophages and by epithelia of the skin and intestine. It requires proteolytic activation, usually via inflammasome-dependent caspase-1 activation, and is thought to escape the cytosol through Gasdermin-D-mediated pyroptosis. Active IL-18 has a high affinity, soluble inhibitor called IL-18 Binding Protein (IL-18BP), which is itself induced by interferon-gamma (IFNg) signaling^8^. IL-18 canonically acts on NK- and activated T-cells to promote type 1 cytokines production and granule-mediated cytotoxicity. It usually acts in concert with cytokines of the Jak-STAT pathway like IL-12 or IL-15. However, it may serve homeostatic roles at tissue sites. Reports of serum IL-18 elevation abound across a wide variety of infectious, malignant, and rheumatic diseases. However, the extraordinarily high serum levels necessary to overcome inhibition by IL-18BP and generate “free IL-18” appear restricted to diseases at highest risk of Macrophage Activation Syndrome (MAS), including Systemic Juvenile Idiopathic Arthritis (SJIA), Adult-Onset Still’s Disease (AOSD), and monogenic diseases like NLRC4-MAS and C-terminal mutations in the Rho GTPase CDC42^8-11^.

## METHODS

Patients were recruited to, consented to, and evaluated as part of Natural History protocols approved by the Institutional Review Boards of 1) the intramural research program (IRP) of the National Human Genome Research Institute, 2) the IRP of the National Institute of Allergy and Infectious Diseases, 3) the University of Pittsburgh, and 4) the Hospital Clinic/Institut d’Investigacions Biomèdiques, Barcelona. All patients Patients with PAPA were defined has having at least one of the cardinal symptoms (pyogenic arthritis, pyoderma gangrenosum, and severe acne/hidradenitis suppurativa) in the presence of a known mutation in *PSTPIP1*. “PAPA-like” patients had at least one of these symptoms in the absence of such a mutation. Serum total IL-18, IL-18BP, and CXCL9 were measured as in Weiss et al.^8^. Briefly, serum was diluted 25-fold and assayed on a Magpix or FLEXMAP 3D multiplex instrument per the manufacturer’s instructions (Luminex). Recombinant IL-18 and CXCL9 were used as standard (MBL International and Peprotech, respectively). Human IL-18BPa-Fc (R&D Systems) was run separately given its interaction with recombinant IL-18^8^. IL-18 and IL-18BPa beads were generated by conjugating capture antibody to magnetic beads per the manufacturer’s instructions (Bio-Rad), whereas CXCL9 beads were purchased (Bio-Rad). Minimal variation between plates and runs was verified using bridging controls. Free IL-18 was measured as previously demonstrated^11^.

## RESULTS

Twenty-nine patients were identified as having symptoms suggestive of PAPA. Nineteen patients were found to have heterozygous known mutations in *PSTPIP1*, with the p.Ala230Thr (10/19) and p.Glu250Gln (6/19) as the most prevalent (Supplemental Table 1). By contrast, 10 PAPA-like patients did not carry such mutations in *PSTPIP1*. One PAPA-like patient (pyoderma gangrenosum) carried the Arg405Cys variant, whereas a patient with an undifferentiated AID (recurrent fevers) carried a Gly258Ala variant, both of which we classified as benign.

Clinical features overlapped substantially between patients with and without *PSTPIP1* mutations, consistent with their pattern of referral. However, patients harboring *PSTPIP1* mutations were younger at disease onset, more had a history of arthritis, and fewer had had pyoderma gangrenosum (Table 1). Both groups’ treatment history reflected the often-recalcitrant nature of these symptoms, with many patients having been treated with glucocorticoids and more than one biologic medicine (Supplemental Table 1). Two patients had clinical features of PAPA but also cytopenias, organomegaly, and mutations associated with the Hyperzincemia/hypercalprotectinemia (Hz/Hc) syndrome^12^. No PAPA or PAPA-like patients had a history of MAS. Review of records revealed ferritin measurements of at least one timepoint in 10 PAPA and 8 PAPA-like patients, with no instances of hyperferritinemia.

**Table 1:** Clinical Characteristics of PAPA & PAPA-like patients.

| Feature (n) | PAPA (19) | PAPA-like (10) | Univariate p |
| --- | --- | --- | --- |
| Sex (F/M) | 9/10 | 6/4 | n.s.<sup>a</sup> |
| Age at onset Median (range) | 1.25 (0.17 to 45) | 12.5 (3 to 19) | p=0.038<sup>b</sup> |
| Arthritis (y/n) | 18/1 | 2/8 | p<0.0001<sup>a</sup> |
| Cystic Acne# | 1 (0 to 3) | 2 (0 to 3) | n.s.<sup>c</sup> |
| Pyoderma Gangrenosum# | 1 (0 to 3) | 3 (2 to 3) | p=0.0046<sup>c</sup> |
| Max CRP* (mg/dL) | 2.4 (0.2 to 21.2) | 1.1 (0.4 to 7.2) | p=0.103<sup>b</sup> |

#Median (range) of ordinal scale: 0=never, 1=mild, 2=moderate, 3=severe
\*Median (range) of records available for timepoints associated with biomarker measurements

As part of an effort to determine the distribution of serum IL-18 across autoinflammatory diseases^8^, stored serum from enrolled patients were assayed retrospectively and compared with relevant disease controls. These controls included patients with activating mutations causing NLRC4 inflammasome hyperactivity and chronically-elevated serum IL-18 (NLRC4-MAS) as well as patients with the pyrin inflammasomopathy FMF. Previous work demonstrated that dramatically elevated serum IL-18, and detectable free IL-18, were unique to patients at significant risk for MAS and not for other inflammasomopathies, type I IFN-mediated diseases, or other autoinflammatory diseases^8^. Contradicting this, we observed highly elevated serum IL-18 in patients with clinical symptoms suggestive of PAPA only if they harbored mutations in *PSTPIP1* (Figure 1A). PAPA-like patients’ serum IL-18 was largely in the normal range. As expected, NLRC4-MAS serum was highly elevated. FMF patients’ serum showed a bimodal pattern that straddled normal and modest elevation and did not correlate with genotype (Figure 1A and data not shown). CXCL9 levels were not consistently elevated in any group. Though IL-18BP levels were significantly higher in PAPA than PAPA-like patients, we were nevertheless able to detect free IL-18 in all samples from PAPA patients.

**Figure 1:**
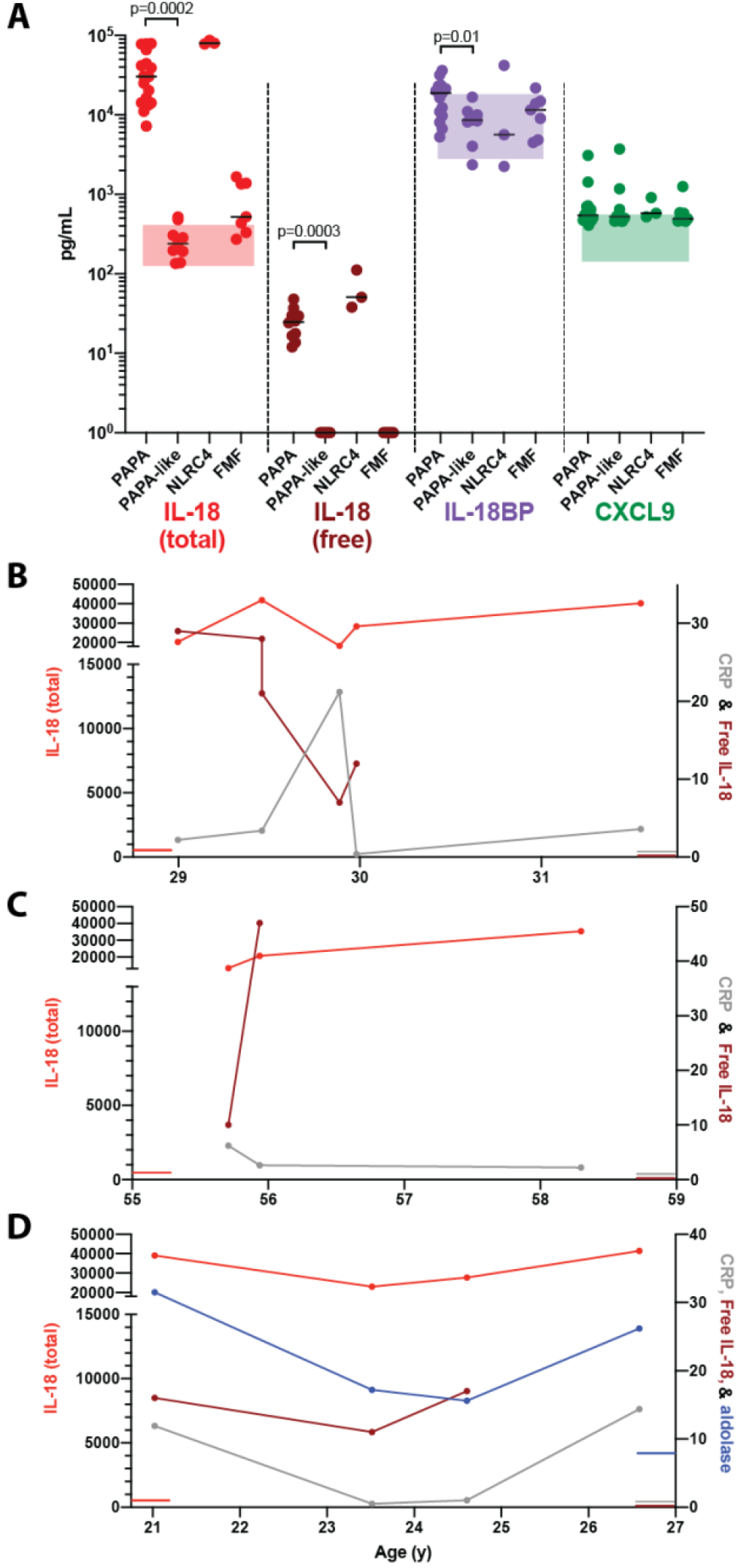
PAPA Syndrome is associated with elevated Total and Free IL-18. A) Serum was assayed for the indicated cytokines. Each point represents the first available sample from each patient. Shaded bars indicate normal range. PAPA and PAPA-like groups were compared by unpaired t-test (displayed) if Benjamini, Krieger, Yekutieli False Discovery Rate <1%. B-D) Longitudinal measurements of IL-18, free IL-18 and C-reactive protein (CRP) from three PAPA patients bearing *PSTPIP1* mutations. Colored lines at y-axes represent upper limits of normal. Total and free IL-18 are in pg/mL, CRP in mg/dL, and aldolase in U/L.

In several patients we were also able to analyze serial samples. Though we observed some variation in total and free IL-18 levels, the degree of this variation was minor in comparison to the degree of C-reactive protein variation (Figures 1B, C, and D). All patients had dramatic elevation of total IL-18, and detectable free IL-18, at all timepoints. ALT and platelet count were not routinely available but when measured were normal even at the peak of CRP elevation (data not shown). Serial aldolase measurements were available in one patient and trended with CRP (Figure 1D).

## DISCUSSION

Many infectious, oncologic, or rheumatic causes of systemic inflammation have been associated with elevated peripheral levels of IL-18, and even very small differences have been independently associated with worse outcomes in chronic inflammatory diseases like atherosclerosis^13^. as high as a few thousand pg/mL. However, IL-18 has an extraordinary dynamic range of over 3 logs in human serum, and extremely high total IL-18 levels have heretofore been observed only in diseases associated with MAS, including SJIA, AOSD, and a few rare monogenic immune dysregulation disorders such as NLRC4-MAS^8^. This has spurred investigation of IL-18 as a fundamental cause of MAS and a clinical trial of IL-18 blockade in genetically-mediated MAS is in progress (NCT03113760).

Though PAPA patients do not appear to be at risk for MAS, we found highly and chronically elevated total serum IL-18, and detectable free IL-18, in mutation-positive PAPA patients. IL-18 may be elevated in PAPA syndrome without increasing MAS risk for a variety of reasons. First, although ample animal work and preliminary case reports suggest otherwise^8, 14, 15^, it is possible that IL-18 elevation is associated with but not contributory to the pathology of either disease. Second, IL-18 typically functions by amplifying the effects of other cytokines, and it may operate in a substantially different inflammatory context in PAPA than SJIA- and NLRC4-MAS. Finally, the source of IL-18 may dramatically alter its effects. The sources of extreme and chronic IL-18 elevation remain unclear. Macrophages are the canonical sites of inflammasome activation and IL-18 production, and *PSTPIP1* mutations have been shown to activate the pyrin-inflammasome in macrophages *in vitro*^16^. However, recent work in *Nlrc4*-hyperactive mice suggests (intestinal) epithelial cells have both inflammasome machinery and abundant pro-IL-18 as a substrate^8^. Notably, skin epithelium is a substantial source of *Il18* transcript as well^8, 13^.

Regardless measurement of peripheral IL-18 may be diagnostically useful in the evaluation for PAPA syndrome regardless of its pathogenic role. The difference in IL-18 between *PSTPIP1* mutation-positive and -negative patients appeared binary, and helped confirm the p.Arg405Cys and p.Gly258Ala variants as likely non-pathogenic. Serum was available from only one patient bearing a PAMI-associated mutation and was elevated similarly to other PAPA patients. Some patients in our cohort had almost exclusively cutaneous or articular disease, suggesting that IL-18 elevation correlates with *PSTPIP1* mutations rather than specific phenotypic features. Thus, serum IL-18 appears to reliably distinguish patients carrying true *PSTPIP1* mutations from patients with suspicious clinical findings or rare variants. Unraveling what connects specific autoinflammatory genes with dramatic elevations of S100 proteins, IL-18, aldolase, and possibly Zinc remains an important area of future research^3, 4^.

Although it includes 19 PAPA patients from various institutions, our study was limited by its relatively small size and retrospective nature. Future studies would benefit from multi-center prospective enrollment and concomitant measurement of other PAPA-associated biomarkers (e.g., aldolase, Zinc, S100 proteins). Nevertheless, our observations add a puzzling diversity to the group of disorders characterized by chronic elevation of total and free IL-18, and outline a path for studying the pathogenic effects of IL-18 beyond MAS.

## Data Availability

n/a

