## Supplemental Table 1 for "Excess Serum Interleukin-18 Distinguishes Patients with Pathogenic Mutations in *PSTPIP1*"

**Supplemental Table 1: Treatment and Genetic information**

| **Protocol** | **Diagnosis** | **Treatments** | **mutation*** | alleles in gnomAD |
|---|---|---|---|---|
| Barcelona | PAPA | GC (PO+IA), canakinumab | PSTPIP1: p.Glu250Gln | 0 |
| Barcelona | PAPA | GC (PO), methotrexate | PSTPIP1: p.Glu250Gln | 0 |
| Barcelona | PAPA | GC (PO), methotrexate | PSTPIP1: p.Glu250Gln | 0 |
| Barcelona | PAPA | GC (PO+IA), anakinra | PSTPIP1: p.Ala230Thr | 0 |
| NHGRI | PAPA | GC(IV+PO), anakinra, canakinumab, infliximab, golimumab, certolizumab | PSTPIP1: p.Ala230Thr | 0 |
| NHGRI | PAPA | GC(PO+IV), anakinra, golimumab, canakinumab | PSTPIP1: p.Glu250Gln | 0 |
| NIAID | PAPA | GC(IV+PO), golimumab, canakinumab | PSTPIP1: p.Glu250Gln | 0 |
| NHGRI | PAPA | anakinra, canakinumab, infliximab, | PSTPIP1: p.Ala230Thr | 0 |
| NHGRI | PAPA / Hz/Hc | GC(PO+IV), anakinra, golilmumab, bone marrow transplant | PSTPIP1: p.Glu257Lys | 0 |
| NHGRI | PAPA | GC(PO+IV), golimumab, anakinra | PSTPIP1: p.Ala230Thr | 0 |
| NHGRI | PAPA | anakinra | PSTPIP1: p.Ala230Thr | 0 |
| NHGRI | PAPA | anakinra | PSTPIP1: p.Ala230Thr | 0 |
| NHGRI | PAPA | GC(PO), infliximab | PSTPIP1: p.Ala230Thr | 0 |
| NHGRI | PAPA | canakinumab, adalimumab | PSTPIP1: p.Ala230Thr | 0 |
| NHGRI | PAPA | GC(IV+PO), anakinra,canukinumab, golimumab, ruxolitinib | PSTPIP1: p.Glu257Gly | 0 |
| NHGRI | PAPA | GC(PO), anakinra | PSTPIP1: p.Ala230Thr | 0 |
| NHGRI | PAPA | golimumab, anakinra | PSTPIP1: p.Ala230Thr | 0 |
| NHGRI | PAPA / Hz/Hc | rilonacept | PSTPIP1: p.Glu250Lys | 0 |
| NIAID | PAPA | GC(PO+IV), anakinra, canakinumab, rilonacept, tocilizumab | PSTPIP1: p.Glu250Gln | 0 |
| NHGRI | PAPA-like | GC(PO), golimumab | PSTPIP1: p.Arg405Cys | 158 |
| NHGRI | PAPA-like | GC(PO), golimumab | none | |
| NHGRI | PAPA-like | GC(IV+PO), anakinra, golimumab, canakinumab | none | |
| NHGRI | PAPA-like | GC(PO), adalimumab,anakinra | none | |
| NHGRI | PAPA-like | anakinra, cyclosporine | none | |
| NHGRI | PAPA-like | golimumab, anakinra, ustekinumab | none | |
| NHGRI | PAPA-like | anakinra, infliximab, ustekinumab | none | |
| NHGRI | PAPA-like | GC(PO+IV), secukinumab, canakinumab | none | |
| NHGRI | PAPA-like | GC(PO), anakinra, golimumab, ustekinumab | none | |
| NHGRI | PAPA-like | adalimumab, doxycycline | none | |
| NHGRI | Undifferentiated | anakinra | PSTPIP1: p.Gly258Ala | 2415 |
| NHGRI | FMF | colchicine | MEFV: p.Val726Ala/Lys695Arg | 561/1648 |
| NHGRI | FMF | colchicine | MEFV: p.Val726Ala/Met694Ile | 561/36 |
| NHGRI | FMF | colchicine, anakinra | MEFV: p.Met694Val/Met694Val | 77/77 |
| NHGRI | FMF | anakinra | MEFV: p.Val726Ala/Glu148Gln | 561/17464 |
| NHGRI | FMF | colchicine | MEFV: p.Met694Val/Glu148Gln | 77/17464 |
| NHGRI | FMF | colchicine, anakinra | MEFV: p.Arg761His/Arg761His | 58/58 |
| Pitt | FMF | GC(IV+PO), colchicine, anakinra | MEFV: p.Met694Val/Met694Val | 77/77 |
| Pitt | NLRC4-MAS | GC(IV+PO), anakinra | NLRC4: p.Gln657Leu | 0 |
| NIAID | NLRC4-MAS | GC(IV+PO), anakinra, canakinumab | NLRC4: p.Thr337Ser | 0 |
| NIAID | NLRC4-MAS | GC(IV+PO), anakinra | NLRC4: p.Val341Ala | 0 |

*All *PSTPIP1* and *NLRC4* mutations appeared in heterozygosity. GC=glucocorticoids. PAPA=Pyogenic Arthritis, Pyoderma, Acne; Hz/Hc=Hyperzincemia/hypercalprotectinemia syndrome.
